# Evaluating a Large Language Model’s Ability to Answer Clinicians’ Requests for Evidence Summaries

**DOI:** 10.1101/2024.05.01.24306691

**Authors:** Mallory N. Blasingame, Taneya Y. Koonce, Annette M. Williams, Dario A. Giuse, Jing Su, Poppy A. Krump, Nunzia Bettinsoli Giuse

## Abstract

**Objective:** This study investigated the performance of a generative artificial intelligence (AI) tool using GPT-4 in answering clinical questions in comparison with medical librarians’ gold-standard evidence syntheses.

**Methods:** Questions were extracted from an in-house database of clinical evidence requests previously answered by medical librarians. Questions with multiple parts were subdivided into individual topics. A standardized prompt was developed using the COSTAR framework. Librarians submitted each question into aiChat, an internally-managed chat tool using GPT-4, and recorded the responses. The summaries generated by aiChat were evaluated on whether they contained the critical elements used in the established gold-standard summary of the librarian. A subset of questions was randomly selected for verification of references provided by aiChat.

**Results:** Of the 216 evaluated questions, aiChat’s response was assessed as “correct” for 180 (83.3%) questions, “partially correct” for 35 (16.2%) questions, and “incorrect” for 1 (0.5%) question. No significant differences were observed in question ratings by question category (p=0.39). For a subset of 30% (n=66) of questions, 162 references were provided in the aiChat summaries, and 60 (37%) were confirmed as nonfabricated.

**Conclusions:** Overall, the performance of a generative AI tool was promising. However, many included references could not be independently verified, and attempts were not made to assess whether any additional concepts introduced by aiChat were factually accurate. Thus, we envision this being the first of a series of investigations designed to further our understanding of how current and future versions of generative AI can be used and integrated into medical librarians’ workflow.

## Introduction

Following the public launch of OpenAI’s Chat Generative Pre-Trained Transformer (ChatGPT) in November 2022 [1], much consideration has been given in the academic and popular discourse to the current and anticipated impact of generative artificial intelligence (AI) on a number of professions. Within the health sciences, studies have investigated the ability of generative AI chat tools (including ChatGPT, Google Gemini, and Microsoft Copilot) to respond to patients’ medical inquiries [2,3], answer questions on licensing exams [4], support healthcare education [5], aid with clinical documentation [6], and contribute to academic manuscripts [7], with many studies focused on specific specialty areas [4]. Authors have also explored the potential utility of generative AI to aid with medical librarians’ professional roles, including developing search strategies for systematic reviews [8,9]. However, the performance of generative AI in the critical task of searching and synthesizing knowledge from the medical literature has not yet been fully assessed, particularly in comparison with medical librarians’ expertise in this area.

At the Center for Knowledge Management at Vanderbilt University Medical Center (VUMC), our team of medical librarians has, for over twenty years, provided evidence syntheses of the biomedical literature to respond to clinicians’ complex questions arising from clinical encounters. These questions were gathered initially through rounding with clinical teams and, since 2004, via a message basket service linked within the electronic health record (EHR) to facilitate clinicians’ ability to send requests at the time and place when they most need an answer [10–14]. A previous study found high levels of physician satisfaction with evidence summaries provided by our team [15]. This service requires the librarian to be highly trained and able to quickly search and filter the current available literature on the topic, extract the most salient information needed to answer the question, and prepare a concise but comprehensive narrative synthesis that is returned to the clinician to inform decision-making [16]. Given the ability of generative AI chat tools to quickly produce detailed, fully articulated summaries drawn from a large body of knowledge, evaluating their current performance in responding to clinical questions is critical to understanding how they may eventually be integrated into medical librarians’ workflows.

Some studies assessing generative AI tools’ ability to provide comprehensive and accurate responses to clinical questions have observed that they can produce accurate results [17–20], particularly for less complex requests [17], although variation in results has been observed among different specialties, tasks, and models investigated [4]. Significant limitations have also been observed, including introduction of both minor and major errors via hallucination or misinterpretation [17,21,22], lack of up-to-date information [23], and limited domain-specific content knowledge [24]. However, with ongoing updates and refinement, it is anticipated that these tools will continue to improve, with advancements already observed, for example, in comparisons of GPT-3.5 to GPT-4 [17,25].

Previous studies have evaluated generative AI chat bots’ responses to clinical questions in comparison with a) published practice guidelines [18,26–28], b) objective multiple-choice answers [4], and/or c) assessment by clinical experts’ review [4,17,29–31]. However, no studies, to our knowledge, have yet evaluated generative AI tools’ responses to actual clinical questions that arise from patient healthcare encounters and use medical librarians’ evidence syntheses as a reference standard. Building upon previous research in knowledge acquisition [32,33] and continuing our examination of how AI could aid or eventually transform medical librarians’ work [34–36], this study aimed to investigate the current ability of aiChat [37], a VUMC-managed generative AI tool, to answer individual facets of clinical questions compared to expertly trained librarians when questions are formulated in a standardized manner. Specifically, the study investigated the following questions:

1. How accurate are aiChat’s responses to clinical questions, as compared with medical librarians’ gold-standard evidence syntheses?
2. Is aiChat’s performance significantly affected by question adjudication status?
3. Are there significant differences in aiChat’s performance by question category?
4. What proportion of references included in aiChat responses can be verified to exist?

## Methods

A sample of actual clinical questions received by our team of information specialists via rounding and an evidence information message basket service linked within the electronic health record (EHR) was used to compare the performance of a locally managed generative AI chat tool with librarians’ gold-standard responses. Although these questions were generated by clinicians in response to specific patient cases, they do not include identifiable patient information, and the study was determined to be exempt by the Vanderbilt University Medical Center Institutional Review Board (IRB 240714). As applicable, this study adhered to the JAMA Network Guidance for Reporting Use of AI in Research and Scholarly Publication [38].

### Generative AI Tool

As submission of proprietary data to public-facing generative AI tools is restricted by our medical center policy, we used an organizationally approved, internally managed AI chat tool called aiChat to conduct the study [37]. At the time of the study, aiChat was a Beta version with options to use either OpenAI’s GPT-3.5 or GPT-4 models. Similar to the public version of ChatGPT, aiChat allows users to submit one or more prompts and receive a response in a user-friendly, conversational format.

### Question Pool

An in-house database used to assign, document, and archive clinicians’ evidence requests answered by our team was queried to retrieve all questions received since 2010 [11,12,14,39]. To align with GPT-4’s most recent knowledge cutoff date at the time of the study, we excluded questions received after April 2023. A group of information scientists then determined eligibility of each archived request. The question set was limited to those that addressed a clinician’s information need during the course of patient care (general education questions and patient education requests were excluded). Additionally, questions were excluded if the evidence synthesis response provided by the librarian contained only a list of citations with no narrative synthesis, reported that no answer was found in the literature, or did not include a clear summary of findings to enable comparison with the aiChat response.

For this initial study, we aimed to assess aiChat’s performance when responding to one simple, focused question at a time, with future analyses planned for assessing performance with complex, multi-faceted requests. Therefore, requests containing more than one distinct topic (e.g., both diagnosis and treatment) were broken into separate facets by information scientists in alignment with the methods established by Giuse et al. [40]. Each facet was considered a separate question for the study. In some cases, questions were reworded for clarity or to remove irrelevant information from the requestor’s original message (e.g., details about requested turnaround time). To allow comparison of performance by question type, each question was assigned one of eight distinct categories: Disease Etiology, Diagnostic Procedure, Differential Diagnosis, Disease Description, Disease Complication, Disease Prevention, Disease Prognosis, or Treatment. These categories were adapted from previous analyses of clinical teams’ information needs [40,41]. All included questions and their corresponding metadata (e.g., date question received, turnaround response time requested) were uploaded to a REDCap database [42,43] for further analysis.

### Determining Critical Elements of the Question Responses

The librarians’ original evidence summaries were used as the gold standard for comparison with aiChat’s summaries. To facilitate the comparison, pairs of medical librarians reviewed the original evidence synthesis response for each included question/facet and came to consensus on the concepts that were most critical and necessary to answer each question. For this process, librarians focused on the most pertinent, high-level conclusions, in recognition that there may be wide variation in wording and other elements within narrative summaries that nonetheless reach the same conclusions. These critical elements were copied from the original response and recorded in REDCap alongside the question.

### Prompt Engineering and Submission

Consultation of the literature for current practices for effective prompt engineering revealed no widely accepted, authoritative guidelines. However, researchers have suggested approaches to improve the quality of generative AI’s response, which were consistent with observations from initial testing by our team, such as giving the chat bot a clear role, establishing the context, and defining the expected output in terms of format and audience [44–46]. The COSTAR framework (Context, Objective, Style, Tone, Audience and Response) [47] was selected to guide prompt engineering for this study as it provides specific details to inform the GPT response, including the use of delimiters to specify the input’s distinct components, and incorporates many of the principles recommended in the literature [47–50]. Using the framework, senior members of the team with expertise in librarianship, knowledge acquisition, medicine, and artificial intelligence devised a standardized prompt to submit with each clinical question (Figure 1).

**Figure 1.**
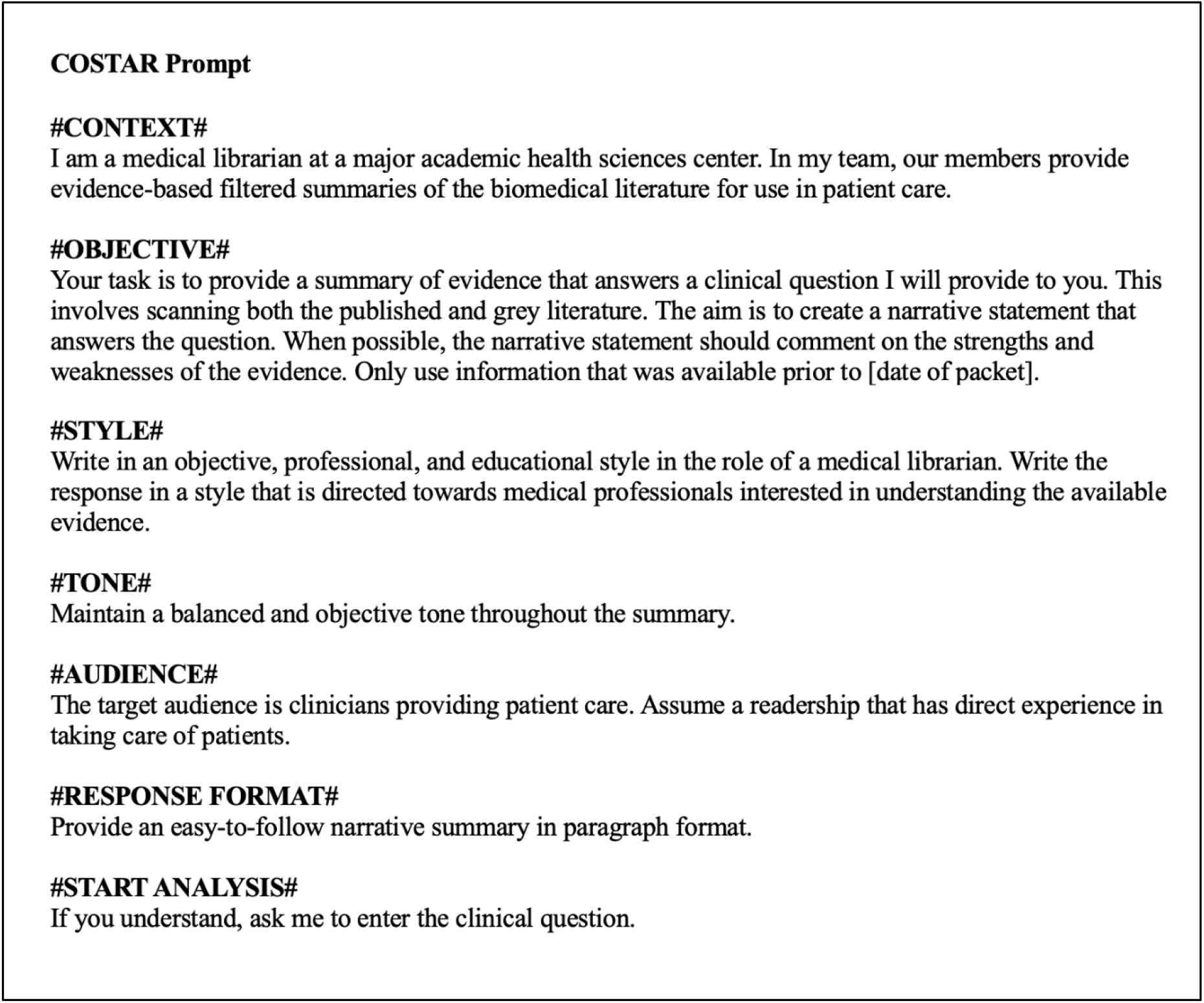
GPT prompt in the COSTAR format.

To avoid inclusion of knowledge to which the librarian would not have had access when originally answering the question, aiChat was prompted to only use information available prior to the date of the original request. In testing, aiChat was able to adjust the response by date when given this parameter. Given that studies have established that GPT often fabricates references [21,51], we did not specifically ask aiChat to provide references as part of the prompt. Providing an example of the desired output within the prompt has also been suggested [50] and found to improve performance in some analyses [9]. However, it is unlikely that a user asking a real clinical question would have an example response readily available to submit, so examples were not included in our prompt.

All questions were submitted to aiChat between March 25, 2024 – April 1, 2024. To capture aiChat’s responses to each question, medical librarians worked in pairs to submit assigned sets of questions to the chat bot tool. First, a librarian selected “New prompt,” set aiChat to use GPT-4, and submitted the prompt (Figure 1). When aiChat responded to confirm understanding (e.g., “Understood. Please enter the clinical question.”), the clinical question was copied directly from the REDCap database and submitted within the same encounter. The full response from aiChat was copied from the interface and saved in REDCap.

Initially, a set of five randomly-selected test questions was submitted to aiChat five times each in sequence by a senior member of our team to assess whether there was enough variation in the responses to necessitate submitting each question multiple times. Although variance was observed in the wording and other elements of aiChat’s summary replies, the overall concepts and conclusions were consistent. Other research has observed significant differences in ChatGPT’s responses when prompts are submitted multiple times [9]. However, the aim of this study was to assess the performance of generative AI for the real-life scenario of a clinician seeking a response to a clinical question. In this context, submitting a question multiple times would not be practical. Thus, for this study, the team decided to submit each question only one time.

### GPT Response Evaluation

Each question, along with the critical elements from the original packet and response from aiChat, was assigned to a pair of medical librarian reviewers for evaluation of the extent to which aiChat’s response aligned with the original librarian’s gold-standard synthesis of evidence from both the published and grey literature. Each reviewer independently assessed whether aiChat answered the question correctly in comparison with the original gold-standard response from the information scientists. The assessment was based on whether aiChat included all, some, or none of the key critical elements that were identified by consensus from the librarian’s original summary. Reviewers used a 3-point Likert scale to indicate whether aiChat’s response was incorrect (1), partially correct or incomplete (2), or correct (3) [31]. The response options avoid the use of non-numerical, vague qualitative terminology (e.g., “mostly correct”) as these types of phrases may create ambiguity and difficulty with interpretation [52,53]. To be considered correct, it was not necessary for aiChat to use the exact same language from the original summary but rather for the response to be conceptually similar. In cases where aiChat provided additional information beyond what the librarian included, the response was not considered incorrect as long as the critical elements were represented.

Discordant ratings were resolved by a third reviewer with medical knowledge and expertise in evidence synthesis, librarianship, knowledge acquisition, and extensive experience with adjudication in knowledge acquisition research [54,55]. The adjudicator thoroughly reviewed each question, the full original summary, the complete aiChat summary, and, if needed, the original supporting references. When relevant, association websites referred to by the aiChat tool were also consulted as of the cutoff date specified in the prompt to confirm whether more recent knowledge may have been incorporated into the response and thus created discrepancies.

### Reference Verification

Although the prompt did not specifically request the inclusion of references, many of aiChat’s responses did include academic references with combinations of author name, journal, and/or publication year. The assessment of accuracy was based only on aiChat’s summary. A separate exploratory analysis was performed using a sub-sample of questions to verify if the references provided by aiChat were real or hallucinated. No attempt was made to compare the references cited with the references selected by the librarian working on the original packet.

For this analysis, a smaller sample of sixty-six questions with citations (30%) was identified through random selection; each question was assigned to a pair of librarians. The librarians reviewed the responses from aiChat and attempted to locate all cited references using the details provided and documented whether the citation was found or not found.

### Statistical Analysis

The ratings for all questions were stored in REDCap and analyzed descriptively using medians, ranges, and frequency. For each question, the absolute (n) and relative frequency (%) of ratings of 1 (incorrect), 2 (partially correct/incomplete), and 3 (correct) was tabulated. The non-parametric Fisher’s exact test, which is powered for data tables where more than 20% of the cells contain value counts of less than five, was used for group comparisons of categorical data. Statistical analyses were conducted using GraphPad Prism 10 software and a two-tailed p-value < 0.05 was used as the threshold for statistical significance.

## Results

The study included 217 discrete questions. During adjudication, one question was excluded due to misclassification as a patient care-related question. The final number of questions analyzed for the study was 216.

Table 1 shows the overall question ratings. Consensus was achieved between librarian pairs on 182 (84.3%) of the questions; the remaining 34 (15.7%) questions required adjudication. Overall, 180 (83.3%) of aiChat responses were assessed as correct in comparison with the original librarian’s response, while 35 (16.2%) were assessed as partially correct and 1 (0.5%) was assessed as incorrect. Results were similar for questions requiring and not requiring adjudication, with 84.1% (n=153) of questions without adjudication and 79.4% (n=27) of questions with adjudication assessed as correct; there were no statistically significant differences in the ratings of questions that received adjudication in comparison to those that did not undergo adjudication (p=0.54). Of the adjudicated questions, most (n=32) were due to a discrepancy of one point (e.g., scores of “2 [partial]” and “3 [correct]”). Two questions were adjudicated due to a discrepancy between “1 (incorrect)” and “3 (correct)” scores.

**Table 1.** Question ratings by adjudication status.

| Questions | 1 (Incorrect) | 2 (Partially Correct/Incomplete) | 3 (Correct) | Total |
| --- | --- | --- | --- | --- |
| Questions without adjudication | 1 (0.5%) | 28 (15.4%) | 153 (84.1%) | 182 (84.3%) |
| Questions with adjudication | 0 (0.0%) | 7 (20.6%) | 27 (79.4%) | 34 (15.7%) |
| **Total** | **1 (0.5%)** | **35 (16.2%)** | **180 (83.3%)** | **216 (100%)** |

### Comparison by Question Category

The most common question category was Treatment (n=147; 68.1%), which included topics such as treatment adverse effects and treatment efficacy, while the least commonly assigned category was Differential Diagnosis (n=1; 0.46%). The percent of aiChat responses assessed as correct was ≥80% across all categories. No significant differences were observed in the question ratings by category (p=0.39). For a full reporting of results by each category, see Table 2.

**Table 2.** Question ratings by category.

| Question Category | Number of Questions | 1 (Incorrect) | 2 (Partially Correct/Incomplete) | 3 (Correct) |
| --- | --- | --- | --- | --- |
| Disease Etiology | 20 | 1 (5.0%) | 1 (5.0%) | 18 (90.0%) |
| Diagnostic Procedure | 10 | 0 (0.0%) | 2 (20.0%) | 8 (80.0%) |
| Differential Diagnosis | 1 | 0 (0.0%) | 0 (0.0%) | 1 (100%) |
| Disease Description | 10 | 0 (0.0%) | 1 (10.0%) | 9 (90.0%) |
| Disease Complication | 8 | 0 (0.0%) | 0 (0.0%) | 8 (100%) |
| Disease Prevention | 7 | 0 (0.0%) | 1 (14.3%) | 6 (85.7%) |
| Disease Prognosis | 13 | 0 (0.0%) | 1 (7.7%) | 12 (92.3%) |
| Treatment* | 147 | 0 (0.0%) | 29 (19.7%) | 118 (80.3%) |
| **Total** | **216** | **1 (0.5%)** | **35 (16.2%)** | **180 (83.3%)** |

*\*aggregates the treatment, treatment adverse effects, and treatment efficacy question categories*

### Comparison by Adjudication and Question Category

The questions sent for adjudication at the highest proportion were related to disease prevention (n=2; 29%); none of the differential diagnosis questions were adjudicated (Table 3). There were no significant differences by category of questions that received adjudication when compared to questions that were not adjudicated (p=0.90).

**Table 3.**
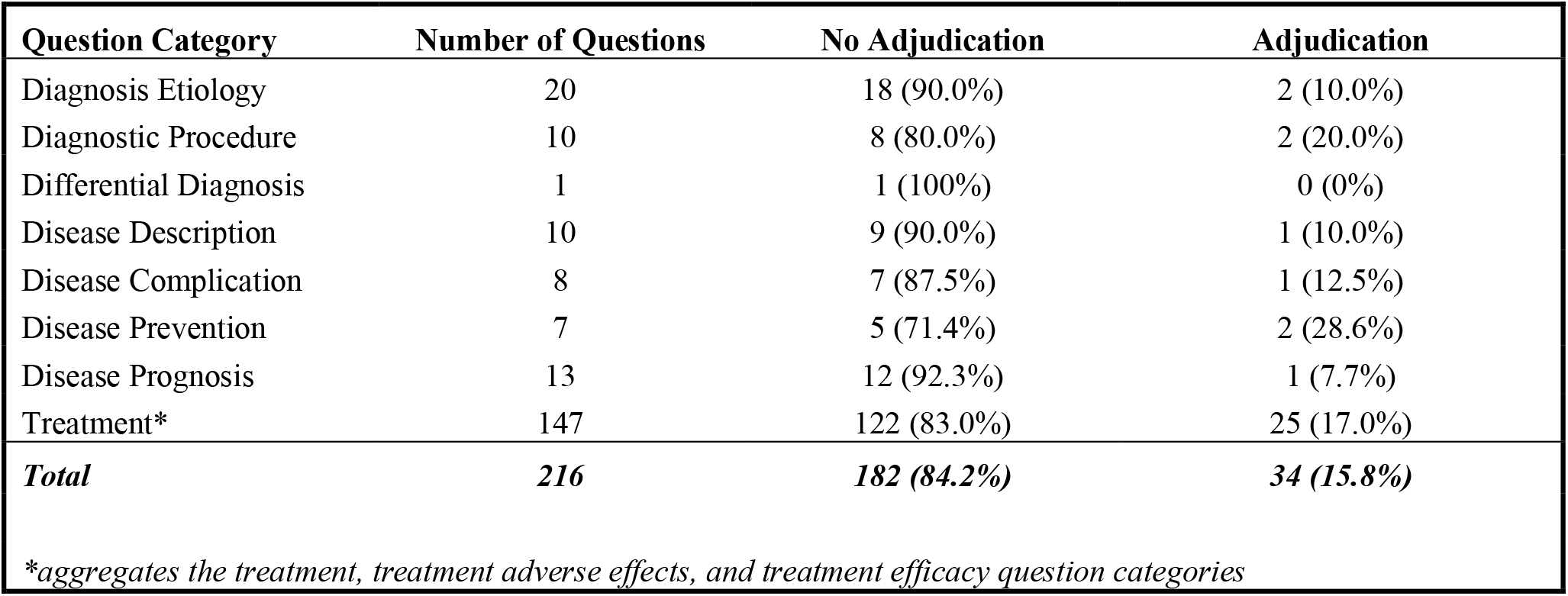
Question adjudication by category.

| Question Category | Number of Questions | No Adjudication | Adjudication |
| --- | --- | --- | --- |
| Diagnosis Etiology | 20 | 18 (90.0%) | 2 (10.0%) |
| Diagnostic Procedure | 10 | 8 (80.0%) | 2 (20.0%) |
| Differential Diagnosis | 1 | 1 (100%) | 0 (0%) |
| Disease Description | 10 | 9 (90.0%) | 1 (10.0%) |
| Disease Complication | 8 | 7 (87.5%) | 1 (12.5%) |
| Disease Prevention | 7 | 5 (71.4%) | 2 (28.6%) |
| Disease Prognosis | 13 | 12 (92.3%) | 1 (7.7%) |
| Treatment* | 147 | 122 (83.0%) | 25 (17.0%) |
| **Total** | **216** | **182 (84.2%)** | **34 (15.8%)** |

*\*aggregates the treatment, treatment adverse effects, and treatment efficacy question categories*

### Verification of References from GPT Response

Out of the 66 questions randomly selected for reference verification there were a total of 162 references. The number of references provided by aiChat per question ranged from 1-4 and the median number of references per question was 2.45. Our team was able to verify the existence of 60 of the 162 references (37.0%). Most of the verifiable citations were indexed in PubMed (n=56; 93.3%), with the remaining available on the cited journal’s website (n=2; 3.3%), a professional organization’s website (n=1;1.67%) and the website of the Food and Drug Administration (n=1;1.67%).

## Discussion

In this initial study comparing generative AI summaries with medical librarians’ gold-standard clinical evidence syntheses in response to individual facets of clinical questions, an organizationally managed generative AI chat tool using GPT-4 was able to report key elements identified in the librarian’s evidence synthesis for the majority of clinical questions examined. These results are promising but only a first step in what we foresee to be a series of many investigations into generative AI tools’ ability to summarize the evidence to answer clinical questions. We recognize the complexity and responsibility of creating a valid, comprehensive, and trustworthy evidence synthesis and are cognizant of many of the issues discussed in an article from Zhang and colleagues, including the need to ensure that large language models are trustworthy, transparent, secure, and avoid perpetuating biases [56].

In our sample of clinical questions, aiChat provided a “correct” response for 83.3% of questions and a “partially correct” response for 16.2%, resulting in an overall 99.5% of questions having at least a “partially correct” response. Most of the questions in our study (80.3%) were treatment-related, which is consistent with the types of questions most frequently asked by clinicians [40,41,57]. No significant differences in accuracy were observed across different categories of clinical questions or adjudication status. The one summary rated by the reviewers as “incorrect” was a response to a question about genetic mutations associated with a particular disease, for which aiChat’s response referenced a different gene than the one reported in the gold-standard evidence packet. This finding could possibly suggest a need to better understand how generative AI tools handle genetic information given the complexity of the field.

While the aiChat- and medical librarian-developed summaries were consistent overall in terms of the key concepts included, many (63%) of the supporting references included in a subsample of aiChat’s responses could not be independently verified. The inability to trust references provided by GPT and, consequently, to be able to verify specific details and results of the studies cited in the responses it provides is currently a significant limitation to its use. However, it is possible that generative AI tools’ performance in this area could improve as we continue to see a rise in open access publishing [56,58,59] and the models are not as limited by subscription paywalls.

We also anticipate that GPT’s performance may improve if provided a curated set of articles selected by a medical librarian upon which to base its response. This approach may also aid in addressing ethical concerns with using large language models, which reflect the social biases and inequities present in the clinical research studies and other content included in their training sets [56,60,61]. By selecting content to provide to the generative AI tool, an effort could be made to ensure that references are representative of diverse populations and as free as possible from bias. Tang et al. [22] conducted a study using ChatGPT and GPT-3.5 in which the generative AI tools were provided with content from Cochrane review abstracts from six clinical areas and prompted to provide four-sentence summaries of the systematic reviews. The study found that, in this context, the summaries included few instances of fabrication; however, errors (e.g., those related to misinterpretation of the content) were still observed. In November 2023, OpenAI introduced a feature allowing users to create custom GPTs through which they can provide their own knowledge (e.g., full-text articles or other written documents) for GPT to use when responding to prompts [62]. At the time of the study, this feature was not available through our organization’s internal generative AI tool, but OpenAI does offer the ability to create custom GPTs at the Enterprise level to enable organizations to leverage this option with proprietary information. Tools harnessing generative AI to search and summarize academic papers using underlying literature databases (e.g., Consensus [63] and Scopus AI [64]) are also becoming available. Additional studies are needed in this area to fully understand current models’ ability to accurately summarize research when provided with selected, full-text source material.

In addition to assessing generative AI tools’ performance relative to that of humans, Shah and colleagues have also emphasized the importance of evaluating the benefits of large language models and considering how they can be leveraged to enhance our work rather than simply replicating it [65]. In this study, we observed that a strength of the aiChat responses was the formatting of the narrative summaries, which typically began with a brief introduction to the topic, followed by a well-organized summary with a balanced representation of the viewpoints found in the literature, and ended with brief conclusions. While the requestor receiving the evidence synthesis may be an expert who is already familiar with the topic, they may also wish to share the summary to educate other members of the team with varying specialties (e.g., pharmacists, nutritionists) or who may be more junior (e.g., medical students). Our team recognizes that the approach of establishing the background at the beginning of the response has educational value in our academic setting and considers the inclusion of all viewpoints in the literature to be a best practice for evidence synthesis [41,66]. The organization used by aiChat to structure the responses also has educational value for our profession as a model that can be applied for instructional purposes to train clinical librarians.

### Limitations

An assumption of this study was that medical librarians’ original evidence syntheses accurately reflected the literature as of the original request date, and that clinicians who received the response trusted and agreed that the supporting evidence provided by the librarian answered their questions. Although we did not independently re-verify the information provided in these evidence syntheses, previous studies have found high levels of physician satisfaction with our team’s evidence services [15].

Similarly, we did not assess the accuracy of every detail of aiChat’s summary but rather focused on whether the most critical elements of the librarian’s original response were present and, for a subset of questions, whether references could be verified to exist. No attempts were made in the course of this study to evaluate whether any additional facts introduced by aiChat were accurate, as the comparison was based on whether the critical elements identified in the librarian’s gold-standard response were included in aiChat’s answer.

Finally, it is possible that aiChat’s performance was impacted by elements of prompt design, such as the lack of example in the prompt or our decision to only submit each question once. However, this is likely to replicate real users’ experiences, as busy professionals may not have the time to review multiple responses or have an example response readily available to provide.

### Conclusions

The findings of this study highlight promising performance of a generative AI tool using GPT-4 for providing responses to individual facets of clinical questions, while also confirming known limitations, such as reference fabrication. Since the aim of this study was to evaluate whether aiChat was able to answer clinical questions with an overall response which included the answer given by our established gold standard, we intentionally did not evaluate any additional conceptual differences in the summaries, as we envision this study being the first in a series of investigations. Additional avenues for future research include exploring generative AI’s ability to respond to questions for which librarians found no answer and evaluating aiChat’s answers to complex clinical questions, i.e., questions containing several facets. Given the current inability to independently verify many of the sources used for the generative AI responses, an important next step will be to conduct a more detailed analysis of the source material. A particular area of interest is to establish a better understanding of the extent to which questions can be answered through freely available open source literature. It will also be critical to understand how generative AI performance may improve when provided with a body of literature curated by expert medical librarians. This model could potentially couple GPT’s strengths in summary generation with librarians’ critical expertise in literature selection and assessment.

## Data Availability Statement

The clinical questions used in this study are not publicly available as the data is institutional proprietary information.

## Author Contributions Statement

Mallory N. Blasingame: Methodology; investigation; visualization; writing—original draft; writing—review and editing. Taneya Y. Koonce: Methodology; investigation; data curation; formal analysis; visualization; writing—original draft; writing—review and editing. Annette M. Williams: Methodology; investigation; data curation; visualization; writing—review and editing. Dario A. Giuse: Methodology; investigation; writing—original draft; writing—review and editing. Jing Su: Methodology; investigation; writing—review and editing. Poppy A. Krump: Methodology; investigation; writing—review and editing. Nunzia Bettinsoli Giuse: Conceptualization; methodology; investigation; formal analysis; visualization; writing—original draft; writing—review and editing; supervision.

## Acknowledgements

The authors would like to acknowledge Spencer DesAutels and Sheila Kusnoor for their review and feedback on the manuscript.

